# From Digital File to Transferable Phantom: A Dual-Centre Evaluation of Multi-contrast Phantom for Standardized PET Quantification

**DOI:** 10.1101/2025.05.30.25328639

**Authors:** Sai Kiran Kumar Nalla, Quentin Maronnier, Tala Palchan-Hazan, Olivier Caselles, John A. Kennedy

**Affiliations:** IUTC - Institut universitaire du cancer de Toulouse, Université de Toulouse, Toulouse, France; Department of Medical Physics, Oncopole Claudius Regaud, IUCT Oncopole, Université de Toulouse, Toulouse, France; Department of Nuclear Medicine, Rambam Health Care Campus, P.O.B. 9602, 3109601 Haifa, Israel; Tachyons SAS, Toulouse, France; Ruth and Bruce Rappaport Faculty of Medicine, Technion Israel Institute of Technology, Haifa, Israel

## Abstract

**Background:** Three-dimensional printing has been used to build cost effective & realistic phantoms but their adaptability across facilities remains under-explored. Validation of standardization protocols and quantitative metrics of positron emission tomography (PET) to printed phantoms has been sparse. This study aims to evaluate the digital transferability of a multi-contrast phantom design and the quantitative consistency of metrics across two independent facilities, as a foundational step towards multi-centre standardization.

**Methods:** Cubic phantom inserts (40 mm) featuring spherical targets (0, 10, 15, and 20 mm), of a previously proven design, were fabricated at two different facilities using 3D printing and positioned within a Jaszczak phantom, filled with radiotracer and soap/water solution. Acquisitions (Day 0, 8) were performed on 5-ring Discovery MI and 32 cm OMNI Legend systems. Quantitative metrics, including CT Hounsfield units (HU), PET recovery coefficients (RC) and PET target-to-background ratios (TBR) were statistically analysed (rank sum tests) to assess the influence of facility, scanner model, and acquisition day.

**Results:** CT images revealed minor differences across facilities and scanners, with notable HU improvement (in emulating water) upon prolonged cube immersion. PET image quality was visually good with respect to desired feature representation. Average TBR_Max_ (mean±std. dev.) were 1.56 ± 0.03 (expected 2.0), 2.35 ± 0.05 (expected 2.5), and 3.10 ± 0.05 (expected 3.33). RC_Max_ and RC_Mean_ ranged from 0.69 – 1.11 and 0.56 - 0.89. No statistical differences were found for TBR_Max_, RC_Mean_, or RC_Max_ (p ≥ 0.07 for Mann Whitney; p ≥ 0.57 for Friedman) and Bland-Altman mean bias < 10%.

**Conclusion:** Our study demonstrates that quantitative PET consistency can be reliably maintained across different facilities, printers, and scanners using 3D-printed phantoms produced from a common digital file. This confirmation of digital transferability offers a crucial and cost-effective framework for standardization, paving the way for more accessible multi-center harmonization studies.

## 1. Background

Quantitative Measurements of molecular level biological processes are key tools for diagnosis of diverse diseases in clinical oncology **[1,2]**. Positron emission tomography (PET) in conjunction with computed tomography (CT) is a widely used modality due its ability to provide both structural and functional information at high quantitative accuracy **[3]**. Consistency and reliability of PET/CT systems is ensured by rigorous quality control (QC) standards provided by the National Electrical Manufacturers Association (NEMA) **[4]**, the European Federation of Organisations for Medical Physics (EFOMP) **[5]**, and the European Association of Nuclear Medicine Research 4 Life (EANM EARL) **[6]** by means of test phantoms. Such standards are used to facilitate harmonization and transferability across facilities. Harmonization is generally focused on standardizing acquisition protocols, reconstruction algorithms and quantitative analysis methods ensuring minimal inter-scanner variability **[7–9]**. Whereas, transferability ensures development of robust phantoms and data reproducibility across facilities **[7,8,10].** Recent advancements in PET/CT systems have enabled more precise imaging especially in terms of spatial resolution **[11,12]**. Additionally, the rate of adoption of artificial intelligence (AI) into clinical workflows is rapidly increasing resulting in faster and accurate diagnostic interpretation **[13,14]**. Some other significant advances include the growing contribution of novel radiotracers and greater application of quantitative PET/CT for personalized medicine and theragnostic **[15,16]**. While existing standards provide a foundation, this fast paced innovation calls for new phantom designs and capabilities to push the limits of available testing techniques. Three dimensional (3D) printing is emerging as a powerful tool to create customizable and anthropomorphic phantoms **[17]**. Furthermore, it allows precise control over geometry and material composition which are essential for transferability. Various 3D printing based studies have confirmed that both lesions and tissue uptakes can be modelled for scanner assessment. One such methodology is the usage of permeable grids which can be filled with radioactive tracer **[18,19]**. The internal structure of the grid based on the porosity level can modulate the activity concentration (AC) thus allowing for the creation of varying radiotracer contrast within a single phantom with single tracer injection **[20]**. Multiple studies in both PET and SPECT imaging confirmed the compatibility of such inserts with existing simulation methods **[18]**. Also, 3D printing has been used to create simulated lesions with heterogeneous uptake, aiding in the development of radiomic metrics to bring them one step closer to clinical implementation **[21]**. Additionally, anthropomorphic phantoms mimicking patient geometries that can aid in testing AI algorithms were also produced using this technology **[22,23]**. A key challenge though for adoption of 3D printing is the lack of multi-facility studies to illustrate the ability to rapidly and cost-effectively distribute 3D printed phantoms. More comprehensive studies especially from molecular imaging perspective are needed to establish standardized protocols and data transferability. Material standardization is paramount, requiring reproducible results across different printers especially the characterization of attenuation properties and radiotracer activity distribution. Geometric accuracy is equally critical, necessitating strict quality control during the printing process to avoid dimensional variations that can impact quantitative measurements. A bigger challenge though for widespread adoption of 3D-printing is lack of universally accepted metrics. The choice and variability in metrics can have significant impact on the interpretation of results especially in multi-centre studies **[24]**. The adaptation of existing standardized uptake value (SUV) thresholds to 3D printed grids can be impractical due to contrast bias provided by the grids to the expected value **[20]**, thus other metrics are to be explored. For harmonization between sites, one can rely on each site to accurately prepare the test phantoms to the prescribed contrasts, or one can ship an active phantom from site to site **[23]**. The former method introduces variability by requiring several activity and/or volume measurements, whereas the latter requires long-lived radioisotopes and special shipping. Printed phantoms lend themselves to having the “same” phantom shipped digitally **[23]**, with pre-determined contrast features.

In this study we perform a dual-facility investigation with phantom design specifications provided by **[20]**. While multi-center harmonization is the ultimate goal, a critical prerequisite is a phantom that can be reliably and accurately reproduced across facilities. Therefore, the main aim of this study was to establish the digital transferability of a custom phantom design and validate the consistency of PET/CT quantitative parameters derived from standard image analysis to establish their utility for 3D printing based studies. We aim to assess that a shared design file and protocol could yield reproducible results, thereby providing the foundation for future, large-scale harmonization studies.

## 2. Methods

### 2.1. 3D Printed Inserts Fabrication, Phantom Preparation and Image Acquisition Protocols

Phantom design specifications from a previous study were used, featuring 40 mm porous cubic inserts with varying solidity (S), pore distance (D_P_), and internal spherical targets (T) **[20]**. The total set contained 36 cubes of which 9 were homogeneous (No Target) and 27 with hot targets. Detailed design parameters, including rationales behind S, D_P_, and T configurations, as well as the insert support structure design, are discussed in **[20]**. A brief explanation is provided in appendix (**Figure A1**). The inserts designed were exported in .STL file format which acted as the common shared file. Both facilities used their own FDM printers: a Prusa MK3S+ Printer (Prusa Research, Prague, Czech Republic) for Facility –1 and a Stratasys F170 (Stratasys, Rehovot, Israel) for Facility – 2, with acrylonitrile butadiene styrene (ABS) as the material with print parameters set independently by each facility.

Phantom preparation (**Figure 1)**, followed an adjusted injection protocol (**supplementary information**) from the reference design **[20]**. The cubes were placed in a cylinder from a Jaszczak phantom and filled with soap and water solution 4-6 hours prior to injection. Two studies corresponding to Day 0 and Day 8 were performed at each facility. [^18^F]FDG ([^18^F]-2-fluoro-2-deoxy-d-glucose) injection on each acquisition day was carried out 45-55 mins prior to scan. Tracer was administered by removing 10-50ml of background solution close to the phantom cap prior and was added back prior to scan. The effective activity (**Table 1**) at first scan was maintained as the base criteria for standardization between facilities. At both facilities, image acquisitions were performed on a 5-ring Discovery MI followed by a 32 cm OMNI Legend (*GE Healthcare, Chicago, IL, USA*). Acquisition parameters were maintained across facilities for each scanner. A helical CT scan using three different slice thickness reconstructions were acquired for each scanner – 2.5mm CT (clinical), 1.25mm CT (high resolution) and a 3.5mm CT for the PET attenuation correction. The clinical CT was the main reconstruction presented in this study. All clinical CT’s were acquired at 120 keV with a 512 × 512 matrix and a 50 cm transverse field of view (pixel spacing 0.98mm) for both scanners. PET acquisition parameters are presented in **Table 1**. All PET acquisitions were carried out across 5 consecutive frames. To equalize for different count rates between scanners, the scan duration for each OMNI acquisitions was scaled based on the effective decay and scan duration used for DMI. The exact scan durations are laid out in **Table 1**.

**Figure 1.**
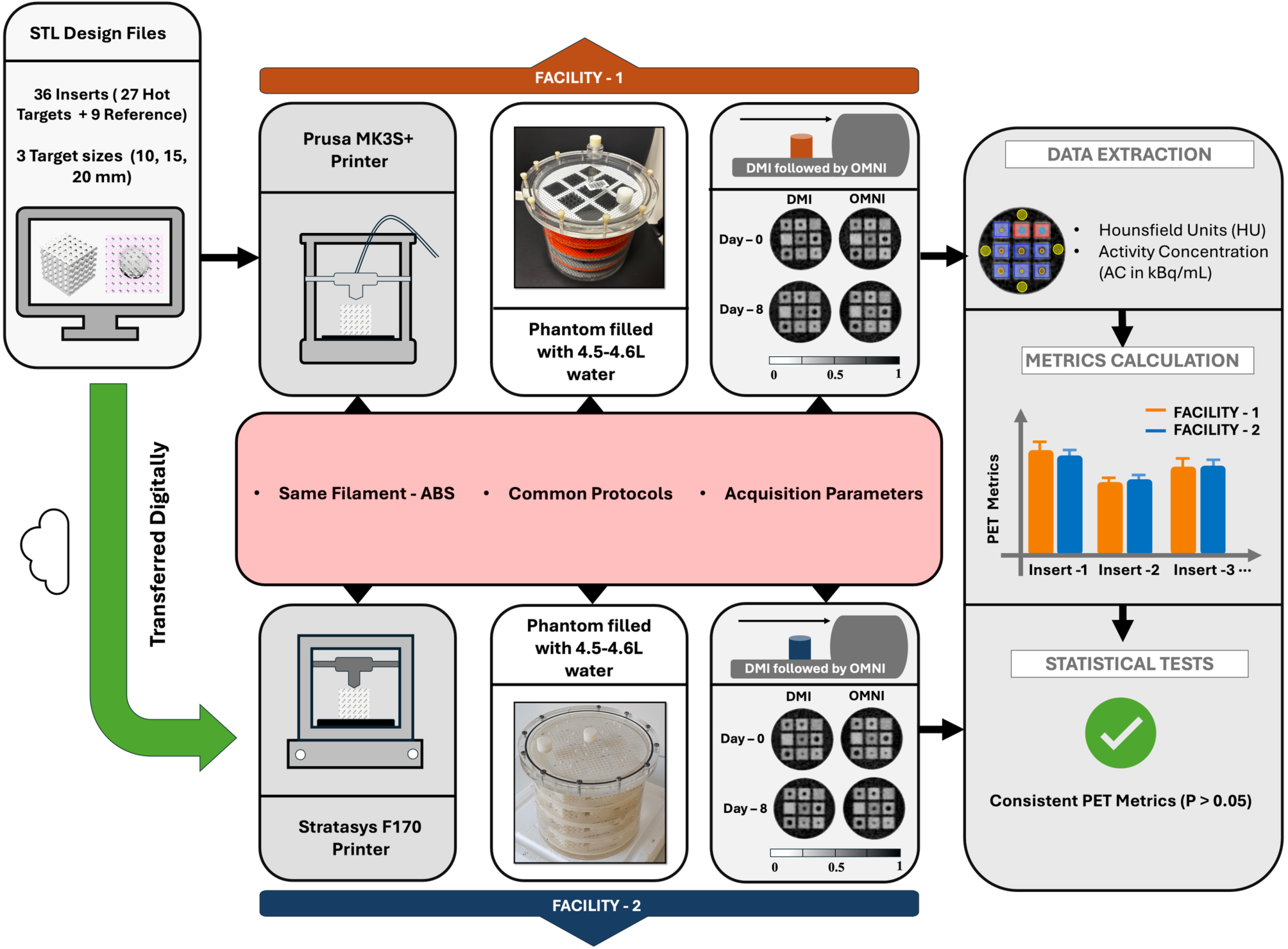
Workflow for dual-facility 3D-printed phantom study. STL design files were printed at two facilities using identical ABS filament and acquisition protocols. (Jaszczak cylinder phantom with 36 cube inserts assembled in 4 layers– exact cube layout for each layer is adapted from **[20])**

**Table 1.** PET Acquisition and reconstruction parameters used by both facilities.

| Parameter | DMI | OMNI |
| --- | --- | --- |
| No.of Bed Positions | 1 | 1 |
| Matrix Size | 256 × 256 |  |
| Slice Thickness | 2.8mm | 2.07mm |
| No. of Slices | 89 | 153 |
| Scan Time | 5 × 300 s | 5 × 420 s |
| Reconstruction Diameter | 600 mm |  |
| Pixel Dimensions | [2.34 2.34] mm |  |
| Voxel Volume | 15.43 mm<sup>3</sup> | 11.33 mm<sup>3</sup> |
| Reconstruction Algorithm | QCFX (time-of-flight BPL*) | QCHD (BPL*) |
| Beta | 500 | 350 |
| Injected Activity in MBq | Facility – 1: 51.0 (*Day 0*), 68.3 (*Day 8*)<br>Facility – 2: 51.1 (*Day 0*), 49.9 (*Day 8*) |  |
| Effective Activity at first scan in MBq | Facility – 1: 37.1 (*Day 0*), 37.1 (*Day 8*)<br>Facility – 2: 37.7 (*Day 0*), 37.5 (*Day 8*) |  |

\*Bayesian penalized-likelihood reconstruction with regularization parameter beta.

### 2.2 Data Extraction and Analysis

Visual comparison of CT and PET Images was performed for qualitative differences. DICOM Images were exported for quantitative data extraction. Hounsfield Units (HU) for CT were extracted manually in 3D Slicer based on VOI workflow used in **[25]**. Quantitative PET data extraction was carried out in MATLAB (*MathWorks INC, MA, USA*) using a semi-automatic program developed in house. Different regions of the image ROIs are in **Figure 2**. Standardized uptake value (SUV), recovery coefficient (RC) & target-background ratio (TBR) were calculated from this data

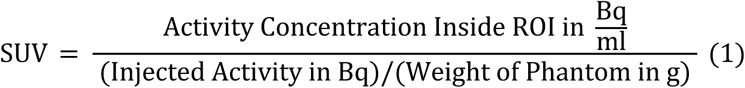

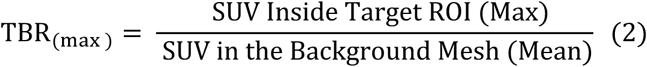

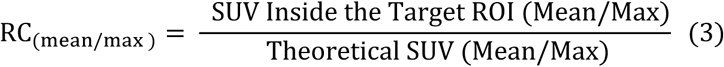

**Figure 2.**
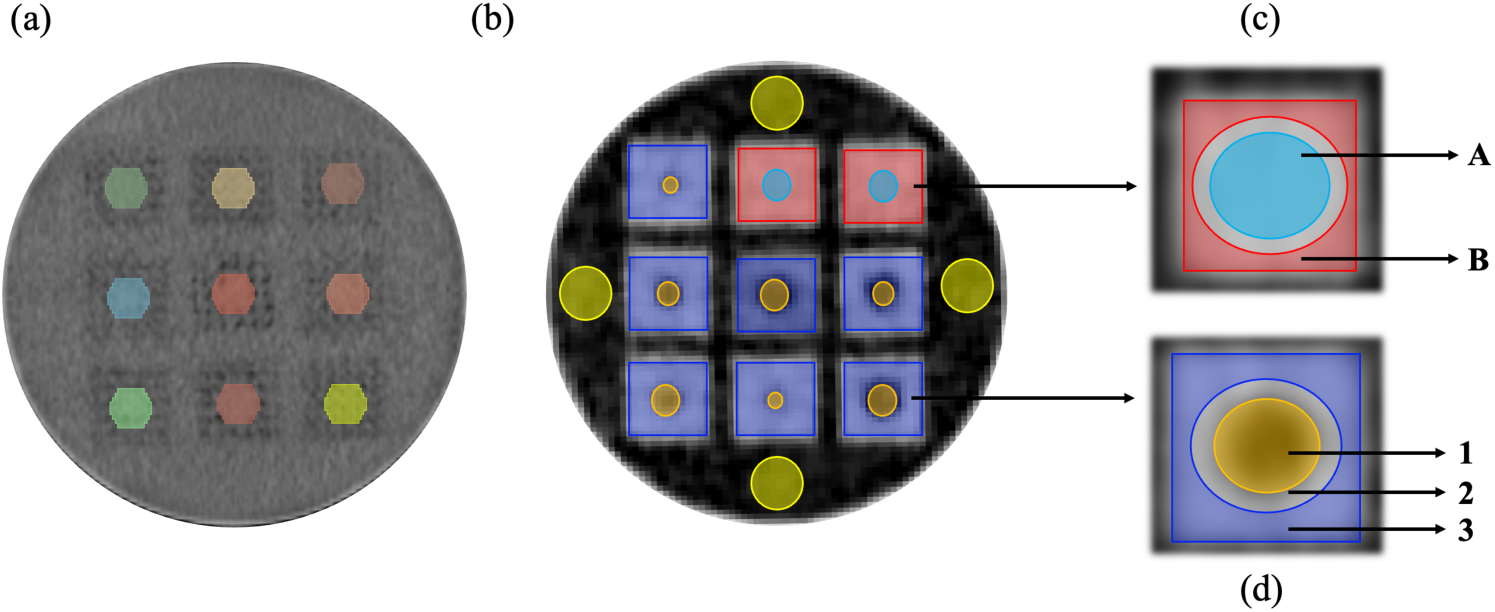
Phantom ROIs. CT: (a) 25 mm Spherical VOI Placement centred on axial plane for HU extraction; (b) ROI placement for each layer on transverse PET slice – background water (yellow), target (orange), homogeneous core (blue), background mesh non target (red) and background mesh target (violet); (c) Exploded view for non-target inserts – central core (region A), corner mesh (region B); (d) Exploded view for target cubes – target (region – 1), no measurement zone (region 2), background mesh (region 3).

Weight of the phantom utilized was 4.5 kg. Data from five consecutive frames were averaged for each acquisition. This study investigated the influence of three key parameters on quantitative results: facility, scanner, and acquisition day. We utilized a hierarchical grouping method to determine the effect of each parameter. For each set, first normality test was performed based on a Shapiro Wilk test followed by non-parametric tests (Mann-Whitney **[25]** , Friedman **[20]**, Wilcoxon signed rank **[26]**). The statistical analyses involved a total of 54 samples for the cross-facility Mann-Whitney U test (27 per facility), 27 paired measurements for pairs tests across the four repeated measures and were performed in JASP (*JASP Team, Netherlands*) and R Studio (*PBC, Boston, MA*) with significance level of 0.05 for the *p values* and ± 10% for the bias and variability.

## 3. Results

### 3.1 CT HU Interpretation

Visually, CT images showed minimal inter-site differences, with quality for all inserts noticeably improving after prolonged immersion (**Figure 3a**). Day 8 results of Facility-1 for D_P_: 5mm are better than Facility -2 with minimal target visibility. Notably the HU_Min_ value within the VOI, revealed the presence of small air bubbles (approximately 2 voxels in size) with very low HU values (-800 to -925) for cubes: 70-5-10 and 70-5-20 on Day 0 (Facility -2), which dissolved over immersion period by Day 8 (**Supplementary Information** - **Figure S2**). Both facilities demonstrated similar initial HU values on Day 0, with a slight increase observed at Facility 1 (-11.89 to -10.68) and a more pronounced increase at Facility 2 (-11.89 to -6.39) by Day 8. Statistical tests yielded *p* values < 0.001 for Shapiro Wilk establishing non-normality also as seen in **Figure 3b**. Mann-Whitney tests resulted in statistically significant differences in HU_Mean_ based on facility (*p value – 0.002*), acquisition day (*p value < 0.001*) and no differences based on scanner (*p value – 0.87*).

**Figure 3.**
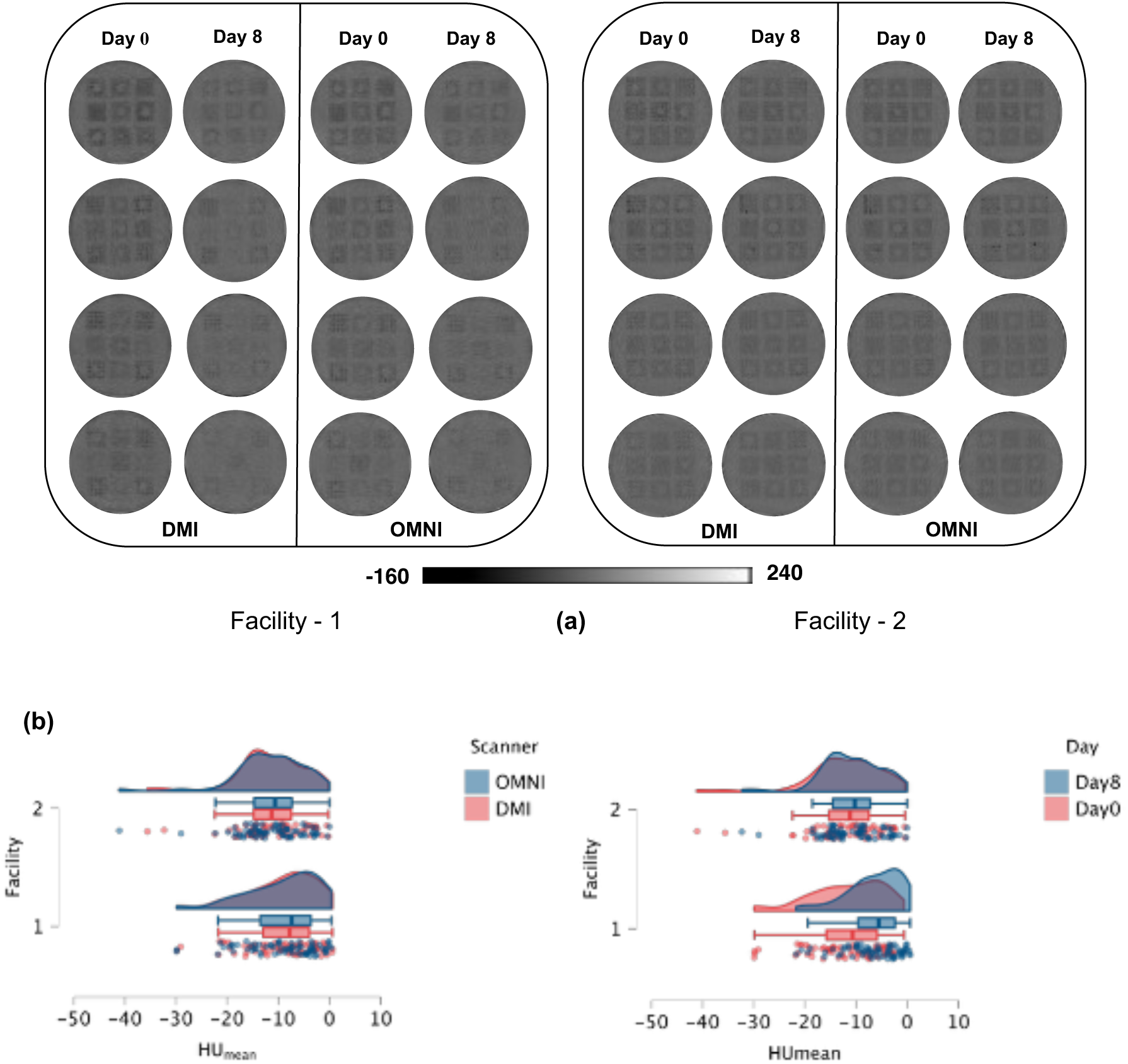
**(a)** CT analysis - Transverse slice of CT for both scanners and facilities viewed on abdominal windowing (W: -40, L: 400; abdominal windowing); **(b)** Kernel Density HU_Mean_ data distribution plots for both facilities separated as function of Scanner (Left) & Acquisition day (Right)

### 3.2 PET Quantitation

For the PET, all slices were able to capture the contrasts and targets visually as illustrated in **Figure 4a**. The TBR and RC are for all days, scanners can be found in **Figure 4b,c (Supplementary Information** – **Table S1)**. The coefficient of variability for noise observed in the images were all less than 5%. SUV variability along the phantom axis **Figure 5a** between layers **[20]** was well within threshold of 10% with maximum variability of 4.1% (Facility - 1) and 8.7% (Facility - 2).

**Figure 4.**
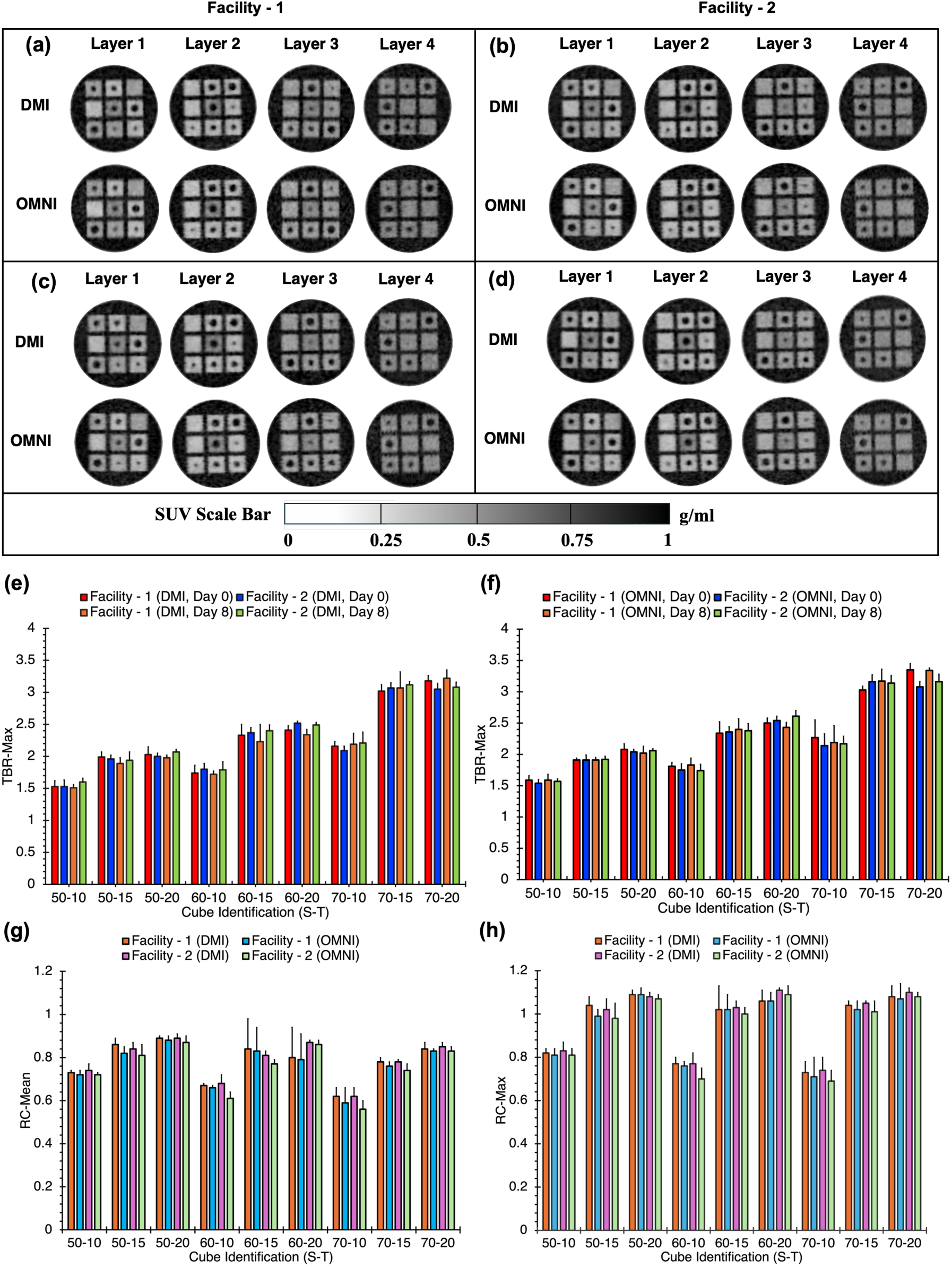
Central Axial Slice of PET for both scanners and facilities: **(a) & (b)** Day 0 and **(c) & (d)** Day 8 *(Note: For Layer -3, the 20mm (3 × 1) and 15mm (3 X 3) cubes of D_P_:10 are misplaced between facilities);* TBR_max_ comparison between facilities (averaged w.r.t unit cell) as a function of S-T for **(e)** DMI and **(f)** OMNI; Recovery coefficient comparison across scanners and facilities for **(g)** RC_mean_ & **(h)** RC_max_

**Figure 5.**
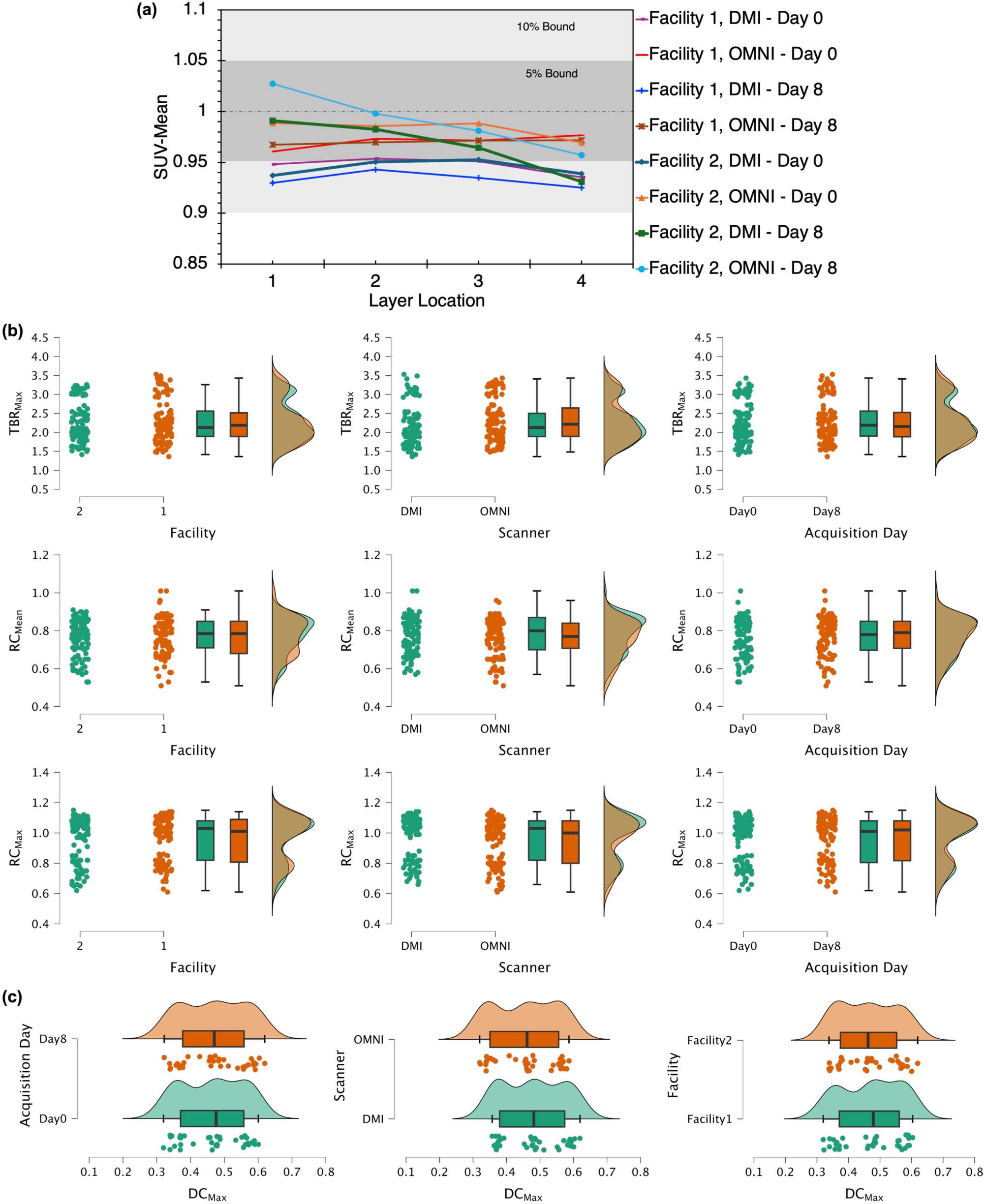
**(a)** SUV in reference solution across layers **(b)** Density Data *overlaid* for 3 quantitative PET metrics of Target Cubes – TBR_Max_, RC_Mean_ and RC_Max_ (Rows) Data assorted together based on Facility, Scanner, and Acquisition Day (Columns); **(c)** Data Distribution Plots for homogeneous cubes –DC_Max_ , Data assorted together based on – Rows: Acquisition Day, Scanner, Facility. (*Expected clusters are centred on 0.3, 0.4, 0.5- A bias in the mean is observed in the acquired data*)

### 3.3 Parametric statistical tests

Statistical Mann Whitney p-values were ≥ 0.07 for all cubes indicating no statistical difference. Additionally, parametric Friedman pairwise analysis resulted p values of ≥ 0.72 (TBR_Max_), ≥ 0.76 (RC_Mean_) and ≥ 0.63 (RC_Max_) for target cubes and ≥ 0.57 (TBR_Max_), ≥ 0.07 (RC_Mean_) and ≥ 0.16 (RC_Max_) for homogeneous cubes. Data distribution of the grouped metrics are represented in **Figure 5b,c**. Both centres observed no statistical differences with all intra facility *p – values > 0.05* and intraclass correlation (ICC) values > 90%. Cross facility statistical results are summarised in **Table 2**. All the metrics have *p >* 0.05 thus no statistical differences for central tendency distribution Bland Altman mean bias fall within the 10% threshold but the SD band width varies for each pair. Post hoc Wilcoxon revealed some outliers with *p < 0.05* (**Table 2**) without alpha rate Bonferroni adjustment.

**Table 2.** Cross Facility Statistical Tests Summary for the PET Metrics of Targets.

| Comparison Pair | Noise (%) | TBR<sub>Max</sub> |  |  | RC<sub>Max</sub> |  |  | RC<sub>Mean</sub> |  |  |
| --- | --- | --- | --- | --- | --- | --- | --- | --- | --- | --- |
|  |  | Mann Whitney | Wilcoxon Bonferroni (True, Adjusted) | Bland Altman | Mann Whitney | Wilcoxon Bonferroni (True, Adjusted) | Bland Altman | Mann Whitney | Wilcoxon Bonferroni (True, Adjusted) | Bland Altman |
| DMI -1<br>vs<br>DMI - 2 | 4.36<br>vs<br>3.60 | 1.00 | 0.16, 0.64 | 2.4<br>(21, -16) | 0.86 | 0.81, 1 | 0.6<br>(11, -10) | 0.55 | 0.05, 0.2 | -1.5<br>(16, -13) |
| OMNI -1<br>vs<br>OMNI - 2 | 4.25<br>vs<br>3.56 | 0.79 | 0.09, 0.36 | -1.4<br>(11, -14) | 0.47 | **0.03**, 0.12 | -1.9<br>(10, -13) | 0.72 | **0.02**, 0.08 | -1.4<br>(13, -16) |
| DMI 1, OMNI 1 correspond to combined data of Facility -1 and DMI 2, OMNI 2 correspond to combined data of Facility – 2<br>Mann Whitney (N<sub>Sample</sub> = 108) and Wilcoxon Sign Rank with Bonferroni Correction (N<sub>Sample</sub> = 108 i.e. 54 pairs) represents the *p values*<br>Bland Altman represents % deviation (Mean Bias, ± 1.96 SD) |  |  |  |  |  |  |  |  |  |  |

## 4. Discussion

We evaluated the standardization and digital transferability of 3D-printed inserts based on the design specifications of **[20]**. This study was carried out across two different facilities, where the shared elements prior to image acquisition were solely the design files (.STL) and the protocols. The quantitative metrics presented a comprehensive assessment of consistency and potential variability in both CT and PET images. Although transaxial CT slices (**Figure 3a**) showed no significant visual differences, Facility 1 demonstrated slightly better HU on Day 8 compared to Facility 2. The change in HU_mean_ values by Day 8 from a common baseline on Day 0 underscores a facility-specific effect, most likely due to printer-related differences. The observed statistical difference in CT HU between facilities (*p* = 0.002), despite using the same material and design file, suggests that inherent printer-to-printer variability can influence the final attenuation properties. While **[20]** identified phantom immersion history as a key factor affecting CT attenuation, our findings highlight its amplified importance in relation to the printer used. This HU change normally stabilizes over a period **[20]** as seen in **Figure 3b** where the range of HU is similar for both facilities on day 8. Although minimal, the presence of small air bubbles within 2 targets on Day 0 for Facility-2 is undesirable. The importance of the chosen immersion strategy is emphasized again, as these bubbles effectively dissipate by Day 8. Additionally the range of HU observed in both facilities are much closer to water / tissue than previous 3D printing studies. Hence, for cross facility studies if equivalent HU_Mean_ is desired, the immersion period needs to be tailored based on material characteristics. Conversely, if the objective is primarily PET quantitation, the effective attenuation correction for 3D-printed grids from CT at 511 keV is accurate across materials **[18,20]** as all the design considerations (D_P_: 5, 7, 10mm) fall well below PET spatial resolution.

PET visual contrasts and target visibility are in agreement with the base phantom design **[20]**. The noise levels within the image are also below the 5% threshold used in standard practice **[5,6]**. SUV variability along the phantom axis based on variability suggested uniform mixing for both facilities. This approach was adopted because the acceptable threshold (10%) **[5]** needed to be consistently achieved regardless of layer location. We adapted the PET preparation protocol to suggestions made by **[20]** by removing and adding back 50 mL volume of the solution during injection and mixing. This change was implemented as the post injection decay time (45-55 minutes) was standardized. This reduced SUV variations in the background and increased reproducibility for the same injected activity.

We observe that the of mean values of achieved TBR_Max_ by 3D printed grids (**Table 2**) are slightly lower than the true values, although they remain within the range defined by the standard deviation (SD). Notably, SD increases with increase in contrast because the percentage of activity inside the mesh of the inserts reduces. This bias which is a function of poisson noise was also previously reported in grid studies **[18–20]**. However, a positive finding is that this bias is consistently captured across facilities and scanners (**Figure 5c and Table S1**). Hence, the potential for a global correction/calibration factor(s) determined based on grid design parameters could be explored. Furthermore, we did not report the TBR_Mean_ for the same reason as it would be unreliable metric. Although the non-significant p-values indicate no detectable statistical differences, the low mean variability (<10%) across contrasts suggests a high level of consistency in TBR_Max_ values, supporting reproducibility. Additionally, the consistent RC values showed no statistical dependence on key parameters and suggest that quantitative results can be reliably reproduced by simply sharing a design file and protocol.

One outlier observed was comparison of RC_Mean_ for homogeneous cubes across scanners which showed significance in Mann Whitney (p ∼ 0.04) but no significance in Friedman (p ∼ 0.07). More specifically, pairwise results of Facility-2 showed inconsistency especially pertaining to a few Day 0 metrics which can be understood in conjunction with presence of air bubbles within 2 targets.

Across facilities, however, the global Mann–Whitney and Friedman tests revealed no significant differences, and the Bland–Altman analysis showed mean biases within the 10% threshold, indicating strong overall agreement between scanners. TBR_Max_ values remained consistent across all pairwise Wilcoxon tests, reinforcing the robustness and transferability of the phantom setup. For recovery coefficients, only two Wilcoxon tests showed statistically acceptable differences, further emphasizing that variability was limited to a small subset of data. Given that the Wilcoxon test is particularly suited to identifying individual outliers, these results mainly highlight a few cubes with potential underperformance rather than widespread inconsistency. To better quantify these deviations, we applied a 10% absolute variability filter, which identified 12% (26 out of 216) of RCMean and 10% (21 out of 216) of RCMax comparisons as outliers.

Though we address the data extraction limitation of **[20]**, the fundamental size of the ROI is still an issue. A key limitation is our ROI definition uses physical dimensions and does not account for printing inaccuracies. An automated segmentation workflow could be better suited for more complex analysis. While we acknowledge the influence of target size on metrics as shown in **Figure 4b,c**, a detailed statistical analysis in this regard was not performed. A key limitation is that cross-scanner pairwise comparison was not performed within scope of this study due to the availability of scanners for experiments. Another limitation of this study is that performance with other radiotracers was not addressed.

The findings of this study emphasize the easy transferability of 3D printed phantoms across facilities with just a single standard design file. The lack of significant differences in PET metrics (RC, TBR) demonstrates that the PET attenuation correction is highly effective at compensating for these small material variations. This finding is critical, as it proves the method is robust and that perfect CT water equivalence is not a prerequisite for accurate PET quantitation with these phantoms. Utilisation of generic low-cost desk FDM printers further enhances the cost-effectiveness of this phantom design with average production cost less than 1000 euros. With the capacity of 36 cubes per acquisition, this opens up the possibility of a wide spectrum of experiments with a single tracer injection. Moving forward, with further studies focusing on wide range of grid design parameters and printers the noise bias in grids could be globally calibrated. Furthermore, exploring post-print processing techniques can reduce and standardize the immersion time needed for CT attenuation stability if necessary. Alternative printing materials, such as PLA, could also lead to improved tissue-equivalent performance on CT **[18]**; however, the immersion vs HU trade off needs an evaluation. This phantom design is intended as a complementary tool rather than a replacement for traditional sphere phantoms. Additionally, a dedicated target detectability study, encompassing a broader range of target sizes (e.g., 5mm to 35mm), is needed to benchmark the performance of these 3D-printed phantoms against established standards. Also a multi-vendor assessment with blind evaluation is required further to find the bottlenecks.

## Conclusion

Our study’s major contribution is the demonstration of robust digital transferability. By distributing only a design file and protocol, we achieved consistency in quantitative PET metrics across two facilities using different printers and scanners. This overcomes a major logistical and financial barrier in multi-center studies: the need to ship heavy, expensive, and sometimes radioactive phantoms. This accessible and low-cost model (average production cost < 1000 euros) can democratize access to high-quality phantom-based quality control.

## Data Availability

All data produced in the present study are available upon reasonable request to the authors

## Financial Statement

NONE

## Data Availability

“*Research data are stored in an institutional repository and will be shared upon request to the corresponding author”*

## Appendix

Porosity (𝜙) or Solidity (S) as parameter to control dilution of radiotracer thus adjust image contrast. The lesion (Target) emulation is achieved through a void and the tissue (Background) emulation with alternating material with controllable design. The dilution is achieved based on below equation

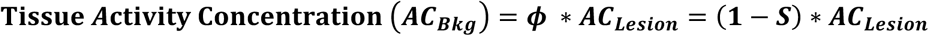

**Figure A1.**
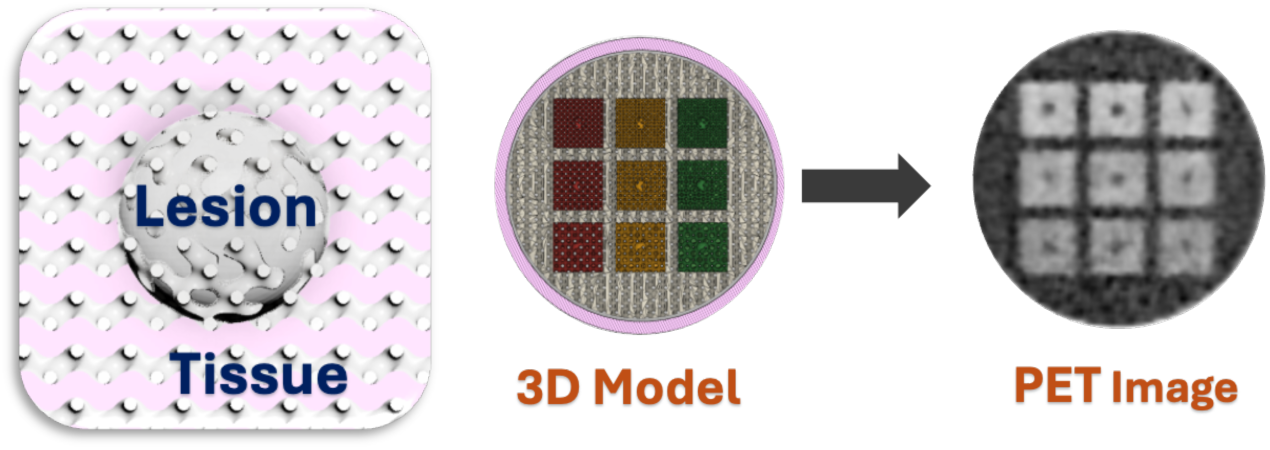
Illustration of the Design

## Data Availability

Data presented in the main article has been included in the supplementary information. Acquired DICOM Images can be made available upon reasonable request to the authors.

## Supplementary Files

### Extended Methods

#### Phantom preparation protocol

The cubes were placed in a cylinder from a Jaszczak phantom and filled with soap and water solution 4-6 hours prior to injection. Two studies corresponding to Day 0 and Day 8 were performed at each facility. Injection on each acquisition day was carried out 45-55 mins prior to scan. Tracer was administered by removing 10-50ml of background solution close to the phantom cap prior and was added back prior to scan.

#### Semiautomatic ROI data extraction

The best transverse slice for each layer was identified automatically for meticulous region of interest (ROI) placement. Exact ROI placement is detailed out in **Figure S1**. Target ROI size is based on converted nearest pixel size from corresponding theoretical dimensions. To mitigate spill-out artefacts and exclude the gradient region where partial volume effects are prominent, a second ROI dilated from target ROI was positioned, and a no measurement zone was established based on Boolean subtraction of the 2 ROI’s. For non-target cubes, 20mm was the reference size for ROI. The extracted activity concentration (kBq/ml) were then converted to standardized uptake value (SUV) with phantom solution weight of 4.5 kg based on Equation 1, with the activities decay corrected to the acquisition start time.

**Figure S1.**
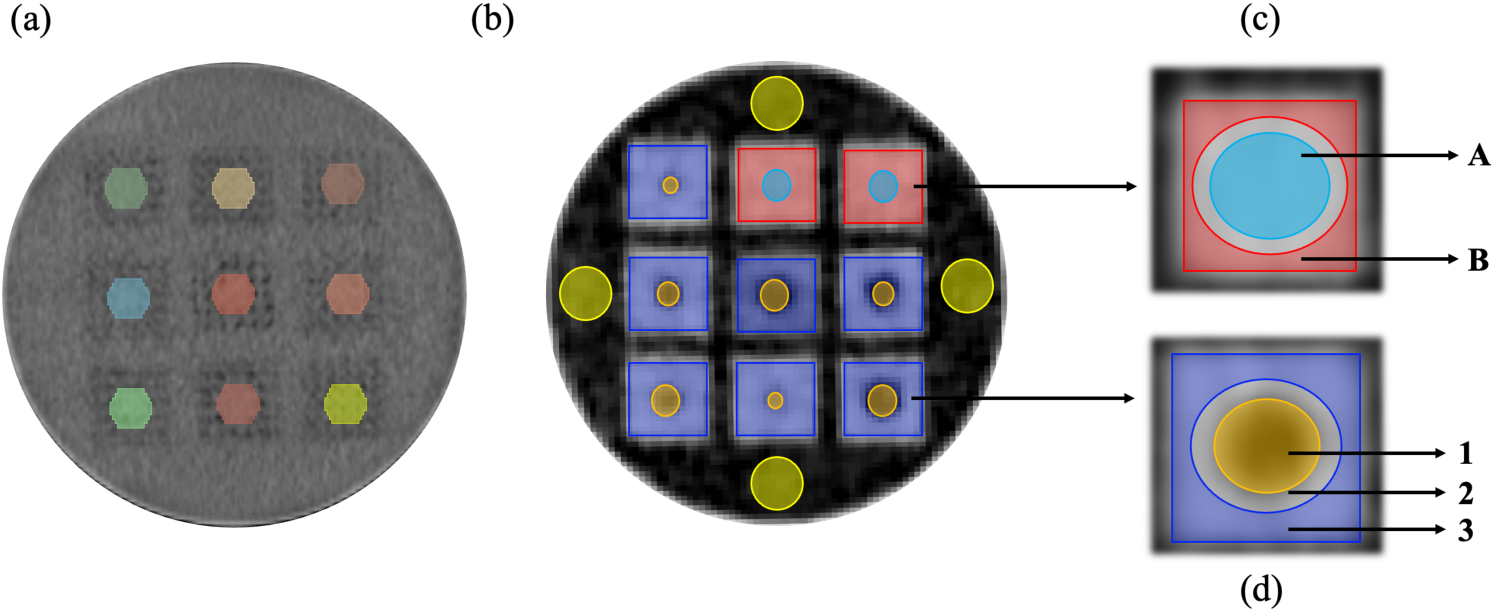
Phantom ROIs. CT: (a) 25 mm Spherical VOI Placement centred on axial plane for HU extraction; (b) ROI placement for each layer on transverse PET slice – background water (yellow), target (orange), homogeneous core (blue), background mesh non target (red) and background mesh target (violet); (c) Exploded view for non-target inserts – central core (region A), corner mesh (region B); (d) Exploded view for target cubes – target (region – 1), no measurement zone (region 2), background mesh (region 3).

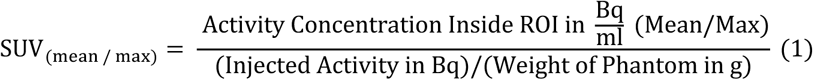

SUV_Mean_ and SUV_Max_ were calculated for the entire data. Background SUV of the solution was averaged over 4 extracted values for each layer. Subsequently to quantify the contrast, target to background ratio (TBR_Max_ and TBR_Mean_) were calculated using equation 2. Expected TBR values based on design parameters (S: 50, 60, 70) are 2, 2.5 and 3.33.

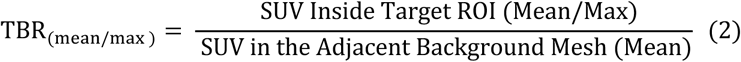

For target quantitation, as per standard practice, recovery coefficient (RC_Max_ and RC_Mean_) were also calculated using Equation 3. Expected value of RC for targets is 1 and for homogeneous cubes is 0.5, 0.4, 0.3 for S: 50, 60, 70.

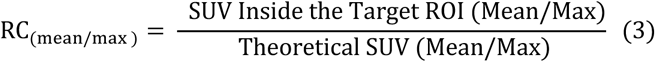

Theoretical SUV is selected as actual measured value in reference solution rather than SUV_theoretical_ ∼ 1, this allows relative assessment of each layer in case of mixing errors. Since all values fall in 10% of SUV_theoretical_ ∼ 1, this should have minimal effect and improves reproducibility.

## Extended Data

**Figure S2.**
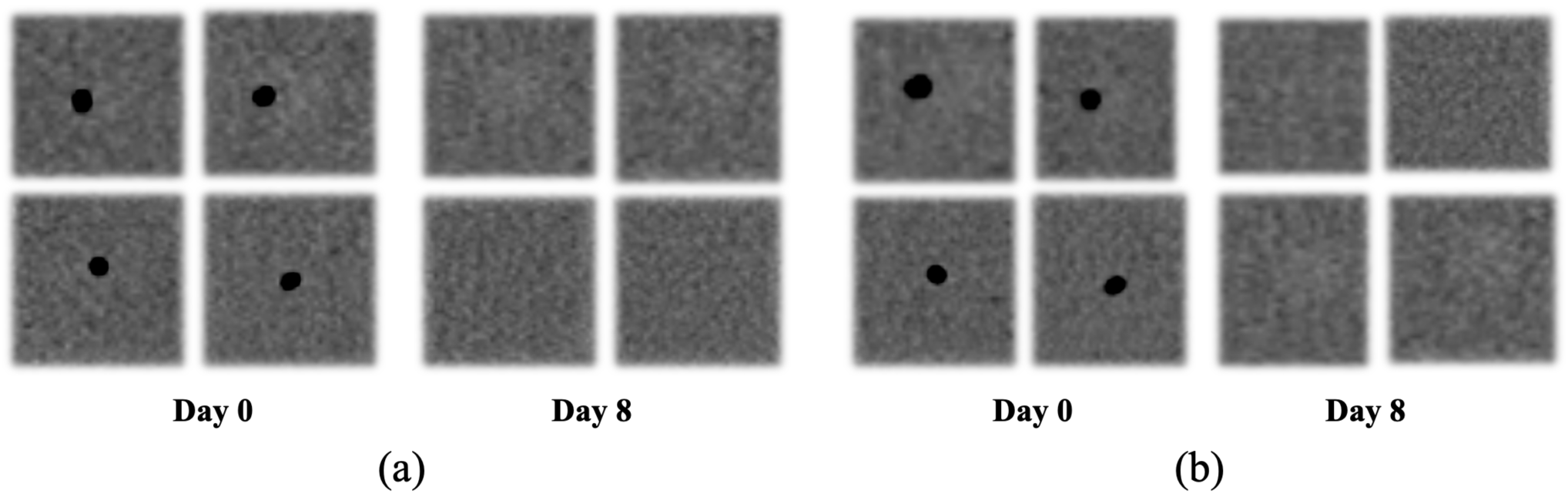
Snapshots of ROI with air bubbles on Day 0 and Day 8 for Facility 2 For DMI at Facility 1, HU value (mean ± Std.dev) was -11.65 ± 14 on Day 0 which changed to -10.8 ± 14 on Day 8. At Facility 2, the initial value was -11.89 ± 14, changing to -6.61 ± 14. OMNI also resulted in same range with the mean values at Facility 1 being -11.89 ± 14 (*Day 0*) and -10.68 ± 14 (*Day 8*) while at Facility 2 they were -11.61 ± 14 (*Day 0*) and -6.31 ± 14 (*Day 8*). For Facility 1, we observed the average noise in the image to be 4.19% for both scanners on Day 0 and 4.54 and 4.3% for DMI and OMNI respectively on Day 8. The average noise values for each scanner respectively at Facility 2 were 3.64 and 3.57% on Day 0, 3.45 and 3.56% on Day 8.

**Table S1.** PET Quantitative Metrics Calculated from Extracted Data.

| Insert Set<br>(Solidity- Target Size) | TBR<sub>Max</sub><br>Day 0 (Mean ± SD); Day 8 (Mean ± SD) | | | |
| --- | --- | --- | --- | --- |
|  | Facility - 1 |  | Facility - 2 |  |
|  | DMI | OMNI | DMI | OMNI |
| 50-10 | 1.53 ± 0.09 | 1.59 ± 0.07 | 1.53 ± 0.10 | 1.54 ± 0.06 |
| | 1.51 ± 0.05 | 1.59 ± 0.09 | 1.60 ± 0.06 | 1.57 ± 0.04 |
| 50-15 | 1.99 ± 0.08 | 1.91 ± 0.03 | 1.96 ± 0.06 | 1.91 ± 0.08 |
| | 1.89 ± 0.09 | 1.91 ± 0.04 | 1.94 ± 0.13 | 1.92 ± 0.05 |
| 50-20 | 2.03 ± 0.12 | 2.08 ± 0.09 | 2.00 ± 0.05 | 2.04 ± 0.04 |
| | 1.98 ± 0.04 | 2.02 ± 0.11 | 2.07 ± 0.04 | 2.06 ± 0.03 |
| 60-10 | 1.74 ± 0.12 | 1.81 ± 0.06 | 1.80 ± 0.09 | 1.75 ± 0.10 |
| | 1.72 ± 0.05 | 1.83 ± 0.11 | 1.79 ± 0.13 | 1.74 ± 0.10 |
| 60-15 | 2.33 ± 0.17 | 2.34 ± 0.18 | 2.37 ± 0.08 | 2.36 ± 0.08 |
| | 2.23 ± 0.27 | 2.40 ± 0.17 | 2.40 ± 0.09 | 2.38 ± 0.11 |
| 60-20 | 2.41 ± 0.07 | 2.50 ± 0.08 | 2.52 ± 0.03 | 2.54 ± 0.07 |
| | 2.34 ± 0.08 | 2.43 ± 0.08 | 2.49 ± 0.04 | 2.61 ± 0.09 |
| 70-10 | 2.16 ± 0.07 | 2.27 ± 0.28 | 2.09 ± 0.07 | 2.14 ± 0.19 |
| | 2.19 ± 0.17 | 2.19 ± 0.27 | 2.21 ± 0.17 | 2.17 ± 0.12 |
| 70-15 | 3.02 ± 0.10 | 3.03 ± 0.06 | 3.07 ± 0.08 | 3.16 ± 0.11 |
| | 3.07 ± 0.25 | 3.17 ± 0.19 | 3.12 ± 0.05 | 3.14 ± 0.12 |
| 70-20 | 3.18 ± 0.08 | 3.35 ± 0.10 | 3.05 ± 0.09 | 3.08 ± 0.08 |
| | 3.22 ± 0.13 | 3.34 ± 0.04 | 3.08 ± 0.08 | 3.16 ± 0.12 |
| Insert Set<br>(Solidity- Target Size) | Averaged RC<sub>Max</sub> (Mean ± SD); RC<sub>Mean</sub> (Mean ± SD) | | | |
|  | Facility - 1 |  | Facility - 2 |  |
|  | DMI | OMNI | DMI | OMNI |
| 50-10 | 0.82 ± 0.02 | 0.81 ± 0.03 | 0.83 ± 0.04 | 0.81 ± 0.03 |
| | 0.73 ± 0.01 | 0.72 ± 0.02 | 0.74 ± 0.03 | 0.72 ± 0.01 |
| 50-15 | 1.04 ± 0.04 | 0.99 ± 0.03 | 1.02 ± 0.05 | 0.98 ± 0.07 |
| | 0.86 ± 0.03 | 0.82 ± 0.03 | 0.84 ± 0.03 | 0.81 ± 0.05 |
| 50-20 | 1.09 ± 0.02 | 1.09 ± 0.03 | 1.08 ± 0.02 | 1.07 ± 0.02 |
| | 0.89 ± 0.01 | 0.88 ± 0.02 | 0.89 ± 0.02 | 0.87 ± 0.03 |
| 60-10 | 0.77 ± 0.03 | 0.76 ± 0.02 | 0.77 ± 0.05 | 0.70 ± 0.05 |
| | 0.67 ± 0.01 | 0.66 ± 0.01 | 0.68 ± 0.04 | 0.61 ± 0.03 |
| 60-15 | 1.02 ± 0.11 | 1.02 ± 0.07 | 1.03 ± 0.03 | 1.00 ± 0.03 |
| | 0.84 ± 0.14 | 0.83 ± 0.11 | 0.81 ± 0.02 | 0.77 ± 0.02 |
| 60-20 | 1.06 ± 0.05 | 1.06 ± 0.04 | 1.11 ± 0.01 | 1.09 ± 0.04 |
| | 0.80 ± 0.14 | 0.79 ± 0.12 | 0.87 ± 0.01 | 0.86 ± 0.02 |
| 70-10 | 0.73 ± 0.05 | 0.71 ± 0.09 | 0.74 ± 0.06 | 0.69 ± 0.05 |
| | 0.62 ± 0.04 | 0.59 ± 0.07 | 0.62 ± 0.04 | 0.56 ± 0.04 |
| 70-15 | 1.04 ± 0.02 | 1.02 ± 0.04 | 1.05 ± 0.01 | 1.01 ± 0.05 |
| | 0.78 ± 0.02 | 0.76 ± 0.02 | 0.78 ± 0.01 | 0.74 ± 0.03 |
| 70-20 | 1.08 ± 0.05 | 1.07 ± 0.07 | 1.10 ± 0.02 | 1.08 ± 0.02 |
| | 0.84 ± 0.03 | 0.83 ± 0.01 | 0.85 ± 0.02 | 0.83 ± 0.02 |

Based on the values in **Table S1,** Cross facility TBR_Max_ (Mean ± SD) for 50% contrast cubes (TBR_Theoretical_ ∼ 2) were 1.82 ± 0.24 and 1.85 ± 0.23 for DMI. The values of same contrast for OMNI were observed to be 1.85 ± 0.21 and 1.84 ± 0.23. Inserts with a TBR_Theoretical_ of 2.5 resulted in values (Facility 1, Facility 2) of 2.13 ± 0.31 and 2.23 ± 0.34 for DMI, 2.22 ± 0.32 and 2.23 ± 0.39 for OMNI. The 70% solidity cubes with TBR_Theoretcial_ of 3.33 yielded DMI values of 2.8 ± 0.5 and 2.77 ± 0.48 while OMNI values were 2.89 ± 0.53 and 2.81 ± 0.5. RC_Max_ and RC_Mean_ were averaged for various insert sets with respect to target sizes for each facility and are presented in **Table S1**.

